# Effects of drinking water, sanitation, handwashing and nutritional interventions on stress physiology, oxidative stress, and epigenetic programming in young children living in rural Bangladesh: A randomized clinical trial

**DOI:** 10.1101/2021.11.24.21266798

**Authors:** Audrie Lin, Andrew N. Mertens, Md. Ziaur Rahman, Sophia Tan, Dora Il’yasova, Ivan Spasojevic, Shahjahan Ali, Christine P. Stewart, Lia C. H. Fernald, Lisa Kim, Liying Yan, Ann Meyer, Md. Rabiul Karim, Sunny Shahriar, Gabrielle Shuman, Benjamin F. Arnold, Alan Hubbard, Syeda Luthfa Famida, Salma Akther, Md. Saheen Hossen, Palash Mutsuddi, Abul K. Shoab, Idan Shalev, Mahbubur Rahman, Leanne Unicomb, Christopher D. Heaney, Patricia Kariger, John M. Colford, Stephen P. Luby, Douglas A. Granger

## Abstract

**Importance:** A regulated stress response is essential for healthy trajectories, but the integrated effects of early childhood environmental and nutritional interventions on stress physiology are unknown.

**Objective:** To assess the effects of a combined nutritional, water, sanitation, and handwashing intervention on physiological stress response, oxidative stress, and DNA methylation.

**Design, Setting, and Participants:** In a trial in rural Bangladesh, we randomized geographical clusters of pregnant women and their in-utero children into either the combined nutritional, water, sanitation, and handwashing intervention or the control group. Physiological stress response, oxidative stress, and methylation levels of 757 children were measured at ages one and two years. Analysis was intention-to-treat.

**Interventions:** The intervention group received combined nutritional counseling and lipid-based nutrient supplements, chlorinated drinking water, upgraded sanitation, and handwashing with soap (N+WSH). The control group did not receive interventions.

**Main Outcomes and Measures:** We measured four isomers of urinary F2-isoprostanes [iPF(2α)-III; 2,3-dinor-iPF(2α)-III; iPF(2α)-VI; 8,12-iso-iPF(2α)-VI] at year one. At year two, we measured pre- and post-stressor concentrations of salivary alpha-amylase and cortisol, overall methylation of the glucocorticoid receptor (*NR3C1*) exon 1F promoter including methylation levels at the nerve growth factor-inducible protein A (NGFI-A) binding site, mean arterial pressure, and resting heart rate.

**Results:** Children in the N+WSH group had lower levels of F2-isoprostanes compared to controls (difference -0.16 to -0.19 log ng/mg of creatinine, *P*<0.01). Compared to the control group, post-stressor cortisol levels were elevated (0.24 log μg/dl; 95% CI, 0.07 to 0.4; *P*<0.01) and the residualized gain score for cortisol was higher (0.06 μg/dl; 95% CI, 0.01 to 0.12; *P*=0.023) in the N+WSH group. Children in the N+WSH group exhibited decreased logit-transformed methylation of the NGFI-A transcription factor binding site (−0.04; 95% CI, -0.08 to 0; *P*=0.037).

**Conclusions and Relevance:** A nutritional, water, sanitation, and handwashing intervention reduced oxidative stress, enhanced hypothalamic-pituitary-adrenocortical axis activity, and reduced methylation levels in a transcription factor binding site of the glucocorticoid receptor gene. A targeted environmental and nutritional intervention affected the set point, reactivity, and regulation of the physiological stress system in early childhood, which may have implications for long-term health and developmental trajectories.

**Trial Registration:** ClinicalTrials.gov NCT01590095

**Key Points:** *Question:* Do intensive nutritional and environmental interventions alter physiological stress response, oxidative stress levels, and epigenetic programming during the first two years of life?

*Findings:* In this cluster randomized clinical trial of 757 children, a combined nutritional, drinking water, sanitation, and handwashing intervention significantly reduced oxidative stress, enhanced the cortisol response, and reduced methylation levels in a transcription factor binding site of the glucocorticoid receptor gene.

*Meaning:* The integrated nutritional, drinking water, sanitation, and handwashing intervention enhanced adaptive responses of the physiological stress system in early childhood, which may have implications for long-term health and developmental trajectories.

## Introduction

Children living in low-income settings often experience recurrent infections and undernutrition due to inadequate water, sanitation, and hygiene infrastructure and food insecurity that may have lasting impacts on their stress response system critical for healthy growth and development. Although several studies have evaluated the effects of psychosocial interventions on the physiological stress system,^1^ a major gap is the lack of experimental studies assessing the effects of physical health interventions on the hypothalamic-pituitary-adrenocortical (HPA) and sympathetic adrenomedullary (SAM) systems and epigenetic programming in early childhood.

Chronic stress, in the form of poverty, may cause irreversible harm if it occurs during the early years of life, a period of rapid growth and development.^2^ During this period of heightened plasticity, the neuroendocrine-immune network develops and adapts in response to exposure to environmental stimuli.^3^ Stressful stimuli shape the set point, reactivity, and regulation of the two primary neuroendocrine axes – the SAM and the HPA systems – and these axes, in turn, regulate the immune system.^3^ Activation of the SAM system leads to increased blood pressure and heart rate and changes in the levels of salivary alpha-amylase, a biomarker of the SAM system.^4,5^

The HPA axis modulates the SAM system through the production of glucocorticoids. Cortisol, a key glucocorticoid, regulates the immune system, growth factors, and neurodevelopment.^6-13^ Cortisol production follows a circadian rhythm that is developed during the first year of life.^14^ Chronic stress disrupts the tight regulation of this circadian rhythm.^15^ An HPA or SAM challenge, such as an acute physical stressor (e.g., vaccination), is typically used to induce and measure cortisol or salivary alpha-amylase reactivity in children.^16^ This artificial induction of reactivity reflects the magnitude of an individual’s HPA or SAM response to a stressor in a naturalistic setting.^17,18^ Glucocorticoids also regulate genes involved in oxidative stress pathways.^19,20^ Oxidative stress results from an imbalance between generation of reactive oxygen species and elimination of them through the antioxidant defense system.^21^ F2-isoprostanes reflect systemic oxidative damage^22,23^ and are associated with infections and neurological damage.^24,25^

Cortisol binds to the glucocorticoid receptor, encoded by the *NR3C1* gene.^6^ Studies suggest associations between stress and epigenetic modulation of the *NR3C1* gene.^6^ During early childhood, the epigenome undergoes dramatic changes. Environmental factors significantly affect these epigenetic processes, which involve the regulation of gene expression through methylation and chromatin modification. Recent studies are beginning to elucidate the degree to which environmental factors during infancy affect developmental programming at the DNA level to determine health outcomes in adulthood. Within the *NR3C1*, increased methylation of CpG sites of a noncanonical nerve growth factor-inducible protein A (NGFI-A) binding site downregulates the expression of the *NR3C1* gene.^26^ Differential methylation of *NR3C1* or the NGFI-A binding site is associated with childhood maltreatment.^27^

Previously, the WASH Benefits trial reported that children receiving a water, sanitation, handwashing, and nutritional intervention experienced better growth and neurodevelopment compared to children in the control group.^28,29^ Here, we evaluated the effects of the intervention on physiological stress response, oxidative stress levels, and DNA methylation of the *NR3C1* gene among young children.

## Methods

### Study design and randomization

The WASH Benefits trial was conducted in rural villages in the Gazipur, Mymensingh, Tangail, and Kishoreganj districts of Bangladesh.^28^ Clusters in this study were defined as eight neighboring eligible households, and eight geographically adjacent clusters formed a block. A one km buffer zone around each cluster was enforced in order to prevent spillover from nearby clusters. Using a random number generator, an investigator at UC Berkeley (B.F.A.) block randomized clusters to the control arm or to one of the six intervention arms: water; sanitation; handwashing; combined water, sanitation, and handwashing (WSH); nutrition, or combined nutrition, water, sanitation, and handwashing (N+WSH). This study only assessed physiological stress, oxidative stress, and DNA methylation in the control and the N+WSH arm.

Since each intervention delivered had visible hardware, participants and outcome assessors were not masked. However, laboratory investigators were masked to group assignments and four researchers (A.L., A.N.M., S.T., and L.K.), following the pre-registered analysis plan, conducted independent masked statistical analyses. Results were only unmasked after replication of masked analyses.

### Ethics

Study protocols were approved by human subjects committees at icddr,b (PR-11063 and PR-14108), the University of California, Berkeley (2011-09-3652 and 2014-07-6561) and Stanford University (25863 and 35583). In addition, icddr,b organized a data safety monitoring committee that oversaw the study. Participants provided written informed consent.

### Participants

Pregnant women in their first or second trimesters and their children were enrolled in the study.^28^ Households that had plans to move in the following year or did not own their own home were excluded in order to minimize loss to follow-up. Households that utilized a water source with high iron were excluded to optimize the effectiveness of the chlorine-based water treatment intervention.

### Procedures

The control group was a passive control group with no study activities. The N+WSH intervention group received a combination of interventions: nutrition intervention [(lipid-based nutrient supplements that included ≥100% of the recommended daily allowance of 12 vitamins and 9 minerals with 9.6 g of fat and 2.6 g of protein daily for children 6–24 months old and age-appropriate maternal and infant nutrition recommendations (pregnancy–24 months)], water (chlorine tablets and safe storage vessels for drinking water), sanitation (child potties, sani-scoop hoes to remove feces, and a double pit latrine for all households), and handwashing (handwashing stations, including soapy water bottles and detergent soap, near the latrine and kitchen). To promote behaviors such as treating drinking water, using latrines, and handwashing, local community health promoters visited enrolled clusters at least once per week during the first 6 months, and subsequently, at least once every 2 weeks.

One year after intervention, using a previously described protocol,^30^ urine samples for oxidative stress analysis were collected in Briggs Pediatric Sterile U-Bags and preserved with 0.1% thimerosal.

Two years after intervention, we collected saliva specimens. In our stress response protocol for cortisol and salivary alpha-amylase measurements, the acute stressor was a venipuncture and caregiver physical separation from the child. Children refrained from ingesting caffeinated products and medicine at least one hour before the venipuncture stress protocol and the child’s mouth was rinsed with drinking water 15-20 minutes prior to the venipuncture. Using SalivaBio Children’s Swabs (Salimetrics), three saliva samples were collected during the stress response protocol (5-8 minutes before stressor onset, 5 minutes after stressor onset, and 20 minutes after stressor onset). Cortisol was measured at two time points: pre-stressor and 20 minutes post-stressor. Salivary alpha-amylase (sAA) was also measured at two time points: pre-stressor and 5 minutes post-stressor. Additional saliva samples for epigenetic analysis were collected in Oragene kits (OGR-575) and shipped at ambient temperature to EpigenDx (Hopkinton, MA) for DNA methylation analysis of the *NR3C1* gene (details in eMethods in Supplement).

At two years, the resting heart rate of participants was measured with a finger pulse oximeter (Nonin 9590 Onyx Vantage) in triplicate, and systolic and diastolic blood pressure were measured with a blood pressure monitor (Omron HBP-1300) in triplicate.

### Outcomes

Analyses were intention-to-treat. We compared the N+WSH arm versus the control arm separately at one year after intervention (median age 14 months) and two years after intervention (median age 28 months). Outcomes included the concentrations of four isomers of F2-isoprostanes [iPF(2α)-III; 2,3-dinor-iPF(2α)-III; iPF(2α)-VI; 8,12-iso-iPF(2α)-VI] measured at one year after intervention. Pre-stressor and post-stressor concentrations of salivary alpha-amylase (sAA) and salivary cortisol were measured at two years after intervention (median age 28 months). The overall methylation level of the glucocorticoid receptor (*NR3C1*) exon 1F promoter and the difference in percentage methylation at NGFI-A transcription factor binding site (CpG site 12) in DNA samples were measured at two years. Systolic and diastolic blood pressure and resting heart rate were measured at two years.

### Oxidative stress biomarker measurements

F2-isoprostane isomers — iPF(2α)-III; 2,3-dinor-iPF(2α)-III; iPF(2α)-VI; 8,12-iso-iPF(2α)-VI — were quantified by liquid chromatography-tandem mass spectroscopy (LC-MS/MS) at Duke University as previously described and optimized for the present study (details in eMethods in Supplement).^23,31^

### Physiological stress and methylation measurements

Pre-stressor and post-stressor sAA and cortisol were measured following ELISA kit protocols at icddr,b. The initial cortisol sample was undiluted, and the initial dilution was 1:200 for sAA (Salimetrics, Carlsbad, CA). Out-of-range specimens were rerun at higher or lower dilutions. The coefficient of variation for sAA and cortisol outcomes was <10%.

Saliva samples that were to be used for the analysis of DNA methylation were collected in Oragene kits (OGR-575) and analyzed by EpigenDx (Hopkinton, MA). EpigenDx performed salivary DNA extraction from Oragene samples, sample bisulfite treatment, PCR amplification, and pyrosequencing and determined percent methylation.^32^ Methylation levels were assessed across the entire glucocorticoid receptor (*NR3C1*) exon 1F promoter (consisting of 39 assayed CpG sites) (details in eMethods in Supplement).^33^

### Statistical analysis

The pre-registered analysis protocol and replication files for the substudy are available (https://osf.io/9573v/). Analyses were conducted using R statistical software version 3.6.1. All biomarker distributions were right-skewed and thus log-transformed. Percentages of methylation were also skewed and therefore logit-transformed.

We used generalized linear models with robust standard errors and two-tailed p-values. The standard errors account for any repeated measures in the clusters. The randomization of assignment to trial arm resulted in balance in the observed covariates across arms so the primary analysis was unadjusted. For each comparison between arms, we also conducted two secondary adjusted analyses: adjusting for child age and sex only and adjusting for child, age, sex, and covariates found to be significantly related to each outcome (likelihood ratio test *P*<0.2). The full list of covariates is included in the footnotes of the tables.

We conducted a pre-specified analysis stratified by sex since biological differences, differential care practices, or other behavioral practices may influence the effect of the N+WSH interventions.

To determine whether missing specimen rates were random, we compared rates of missing specimens across arms and compared characteristics of participants with missing specimens and those with full sets. To account for imbalances in missing outcomes across arms and potential bias due to informative censoring, we repeated the adjusted analysis using inverse probability of censoring weighting (IPCW), using covariates to predict missing outcomes.^34^

The trial was registered at ClinicalTrials.gov (NCT01590095).

## Results

The overall trial assessed 13279 pregnant women for eligibility. 5551 women were enrolled between 31 May 2012 and 7 July 2013 and randomly allocated to one of the intervention or control groups (Figure 1). The target enrollment for the stress substudy was 996 children after one year of intervention and 1021 children after two years of intervention (Figure 1). Stress outcomes were assessed in 688 children (51% female) at age 14.3 (IQR, 12.7–15.6) months, and 760 children (52% female) at age 28.2 (IQR, 27.0–29.6) months (Figure 1). At enrollment, household characteristics were similar across intervention and control arms (Table 1) and comparable to the overall trial (eTable 1 in Supplement).

**Table 1.**
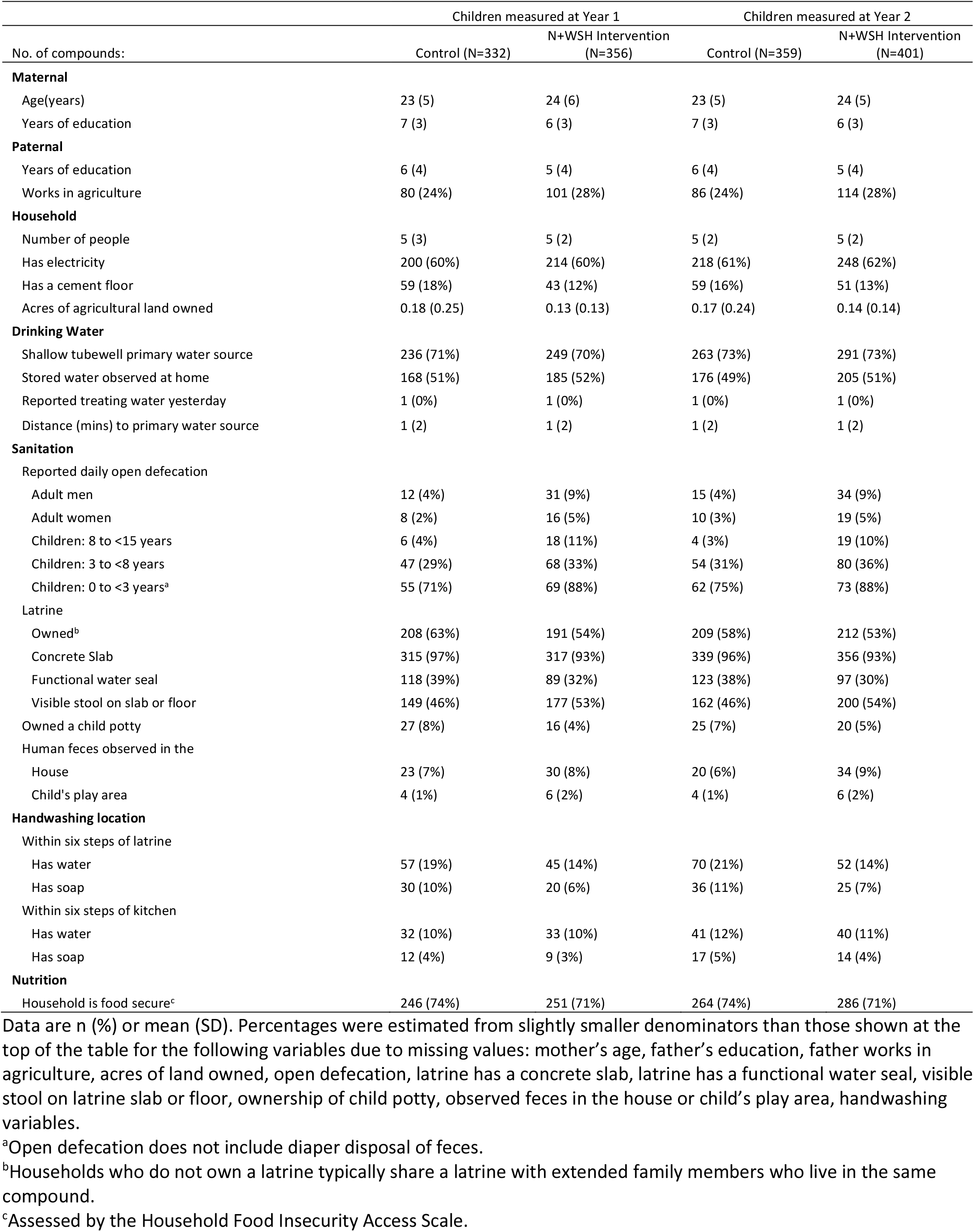
Enrollment characteristics within the Control households and the N + WSH intervention households.

| No. of compounds: | Children measured at Year 1 |  | Children measured at Year 2 |  |
| --- | --- | --- | --- | --- |
|  | Control (N=332) | N+WSH Intervention (N=356) | Control (N=359) | N+WSH Intervention (N=401) |
| **Maternal** |  |  |  |  |
| Age(years) | 23 (5) | 24 (6) | 23 (5) | 24 (5) |
| Years of education | 7 (3) | 6 (3) | 7 (3) | 6 (3) |
| **Paternal** |  |  |  |  |
| Years of education | 6 (4) | 5 (4) | 6 (4) | 5 (4) |
| Works in agriculture | 80 (24%) | 101 (28%) | 86 (24%) | 114 (28%) |
| **Household** |  |  |  |  |
| Number of people | 5 (3) | 5 (2) | 5 (2) | 5 (2) |
| Has electricity | 200 (60%) | 214 (60%) | 218 (61%) | 248 (62%) |
| Has a cement floor | 59 (18%) | 43 (12%) | 59 (16%) | 51 (13%) |
| Acres of agricultural land owned | 0.18 (0.25) | 0.13 (0.13) | 0.17 (0.24) | 0.14 (0.14) |
| **Drinking Water** |  |  |  |  |
| Shallow tubewell primary water source | 236 (71%) | 249 (70%) | 263 (73%) | 291 (73%) |
| Stored water observed at home | 168 (51%) | 185 (52%) | 176 (49%) | 205 (51%) |
| Reported treating water yesterday | 1 (0%) | 1 (0%) | 1 (0%) | 1 (0%) |
| Distance (mins) to primary water source | 1 (2) | 1 (2) | 1 (2) | 1 (2) |
| **Sanitation** |  |  |  |  |
| Reported daily open defecation |  |  |  |  |
| Adult men | 12 (4%) | 31 (9%) | 15 (4%) | 34 (9%) |
| Adult women | 8 (2%) | 16 (5%) | 10 (3%) | 19 (5%) |
| Children: 8 to <15 years | 6 (4%) | 18 (11%) | 4 (3%) | 19 (10%) |
| Children: 3 to <8 years | 47 (29%) | 68 (33%) | 54 (31%) | 80 (36%) |
| Children: 0 to <3 years<sup>a</sup> | 55 (71%) | 69 (88%) | 62 (75%) | 73 (88%) |
| Latrine |  |  |  |  |
| Owned<sup>b</sup> | 208 (63%) | 191 (54%) | 209 (58%) | 212 (53%) |
| Concrete Slab | 315 (97%) | 317 (93%) | 339 (96%) | 356 (93%) |
| Functional water seal | 118 (39%) | 89 (32%) | 123 (38%) | 97 (30%) |
| Visible stool on slab or floor | 149 (46%) | 177 (53%) | 162 (46%) | 200 (54%) |
| Owned a child potty | 27 (8%) | 16 (4%) | 25 (7%) | 20 (5%) |
| Human feces observed in the |  |  |  |  |
| House | 23 (7%) | 30 (8%) | 20 (6%) | 34 (9%) |
| Child's play area | 4 (1%) | 6 (2%) | 4 (1%) | 6 (2%) |
| **Handwashing location** |  |  |  |  |
| Within six steps of latrine |  |  |  |  |
| Has water | 57 (19%) | 45 (14%) | 70 (21%) | 52 (14%) |
| Has soap | 30 (10%) | 20 (6%) | 36 (11%) | 25 (7%) |
| Within six steps of kitchen |  |  |  |  |
| Has water | 32 (10%) | 33 (10%) | 41 (12%) | 40 (11%) |
| Has soap | 12 (4%) | 9 (3%) | 17 (5%) | 14 (4%) |
| **Nutrition** |  |  |  |  |
| Household is food secure<sup>c</sup> | 246 (74%) | 251 (71%) | 264 (74%) | 286 (71%) |

Data are n (%) or mean (SD). Percentages were estimated from slightly smaller denominators than those shown at the top of the table for the following variables due to missing values: mother's age, father's education, father works in agriculture, acres of land owned, open defecation, latrine has a concrete slab, latrine has a functional water seal, visible stool on latrine slab or floor, ownership of child potty, observed feces in the house or child's play area, handwashing variables.
<sup>a</sup>Open defecation does not include diaper disposal of feces.
<sup>b</sup>Households who do not own a latrine typically share a latrine with extended family members who live in the same compound.
<sup>c</sup>Assessed by the Household Food Insecurity Access Scale.

**Figure 1.**
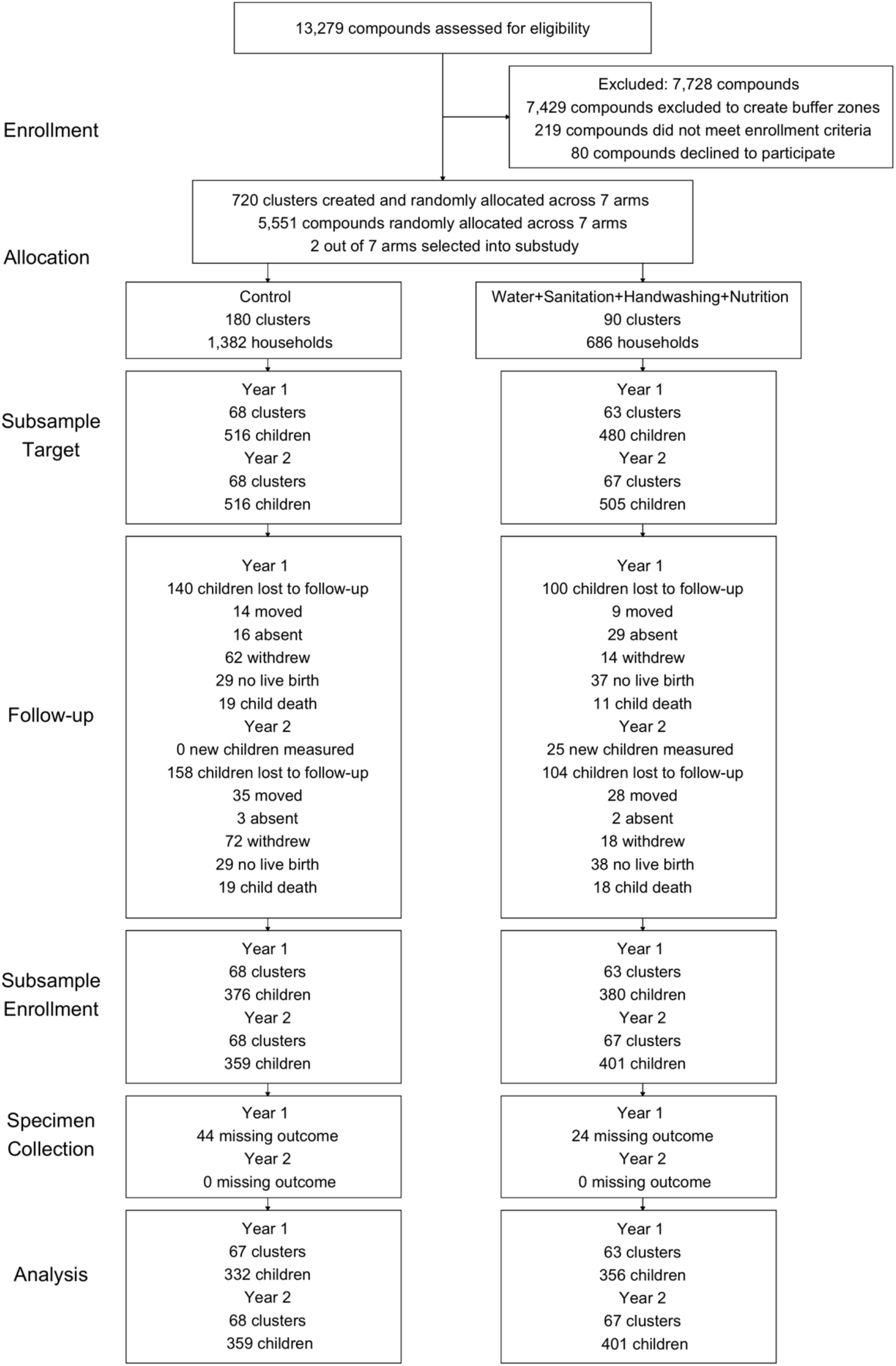
Participant enrollment for the WASH Benefits stress response and DNA methylation study population.

Compared to children in the control group, after one year of intervention (median age 14 months), children in the intervention arm exhibited lower levels of all four F2-isoprostanes isomers measured: IPF(2α)-III (−0.16 log ng/mg of creatinine; CI -0.27 to -0.06, *P*<0.01), 2,3-dinor-iPF(2α)-III (−0.16 log ng/mg of creatinine; CI -0.23 to -0.09, *P*<0.001), iPF(2α)-VI (−0.17 log ng/mg of creatinine; CI -0.25 to -0.1, *P*<0.001), and 8,12-iso-iPF(2α)-VI (−0.19 log ng/mg creatinine; CI -0.29 to -0.1, *P*<0.001) (Figure 2 and eFigure 1 in Supplement; Table 2).

**Table 2.**
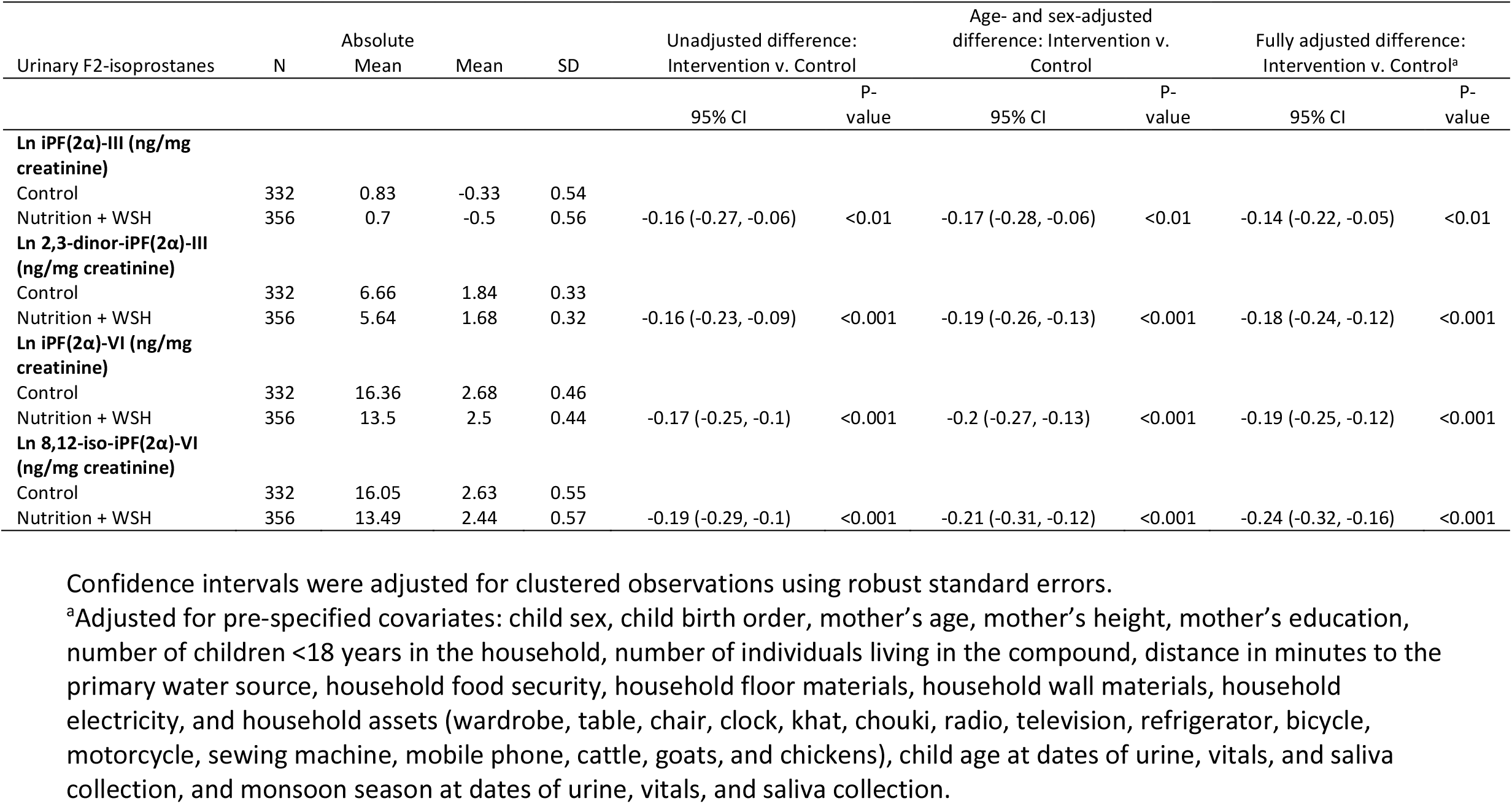
Effect of nutrition, water, sanitation, and handwashing intervention on oxidative stress measurements on Bangladeshi children at age 14 months.

**Figure 2.**
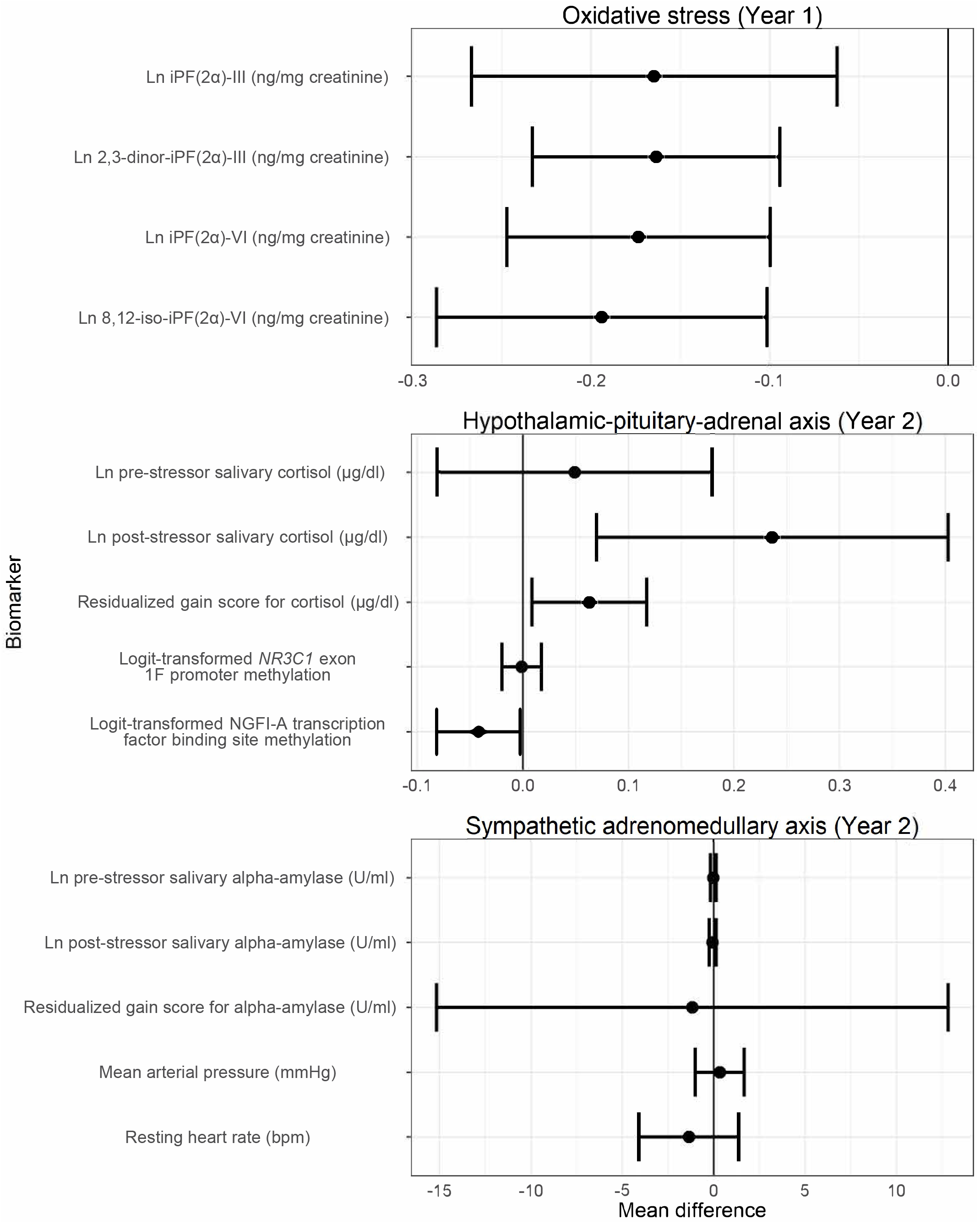
Unadjusted mean differences between the Control arm and the Nutrition + WSH arm for all stress outcomes.

For the cortisol and sAA measurements, the acute stressor was a venipuncture and physical separation of the child from the caregiver. After two years of intervention (median age 28 months), children in the intervention group had elevated post-stressor salivary cortisol levels (0.24 log μg/dl; CI 0.07 to 0.4, *P*<0.01), higher cortisol slope scores (0.002 μg/dl/min; CI 0 to 0.003, *P*=0.035), and higher residualized gain scores for cortisol (0.06 μg/dl; CI 0.01 to 0.12, *P*=0.023; Figure 2 and eFigure 1 in Supplement; Table 3). There was no difference in the overall methylation levels of the *NR3C1* between the control and intervention groups. Logit-transformed methylation of the NGFI-A transcription factor binding site was lower in the intervention group compared to the control group (−0.04, CI -0.08 to 0, *P*=0.037; Figure 2 and eFigure 1 in Supplement; Table 3). Unadjusted, adjusted, and IPCW analyses produced similar estimates (eTables 2 and 3 in Supplement), indicating balance in measured confounders across arms and no differential loss to follow-up.

**Table 3.**
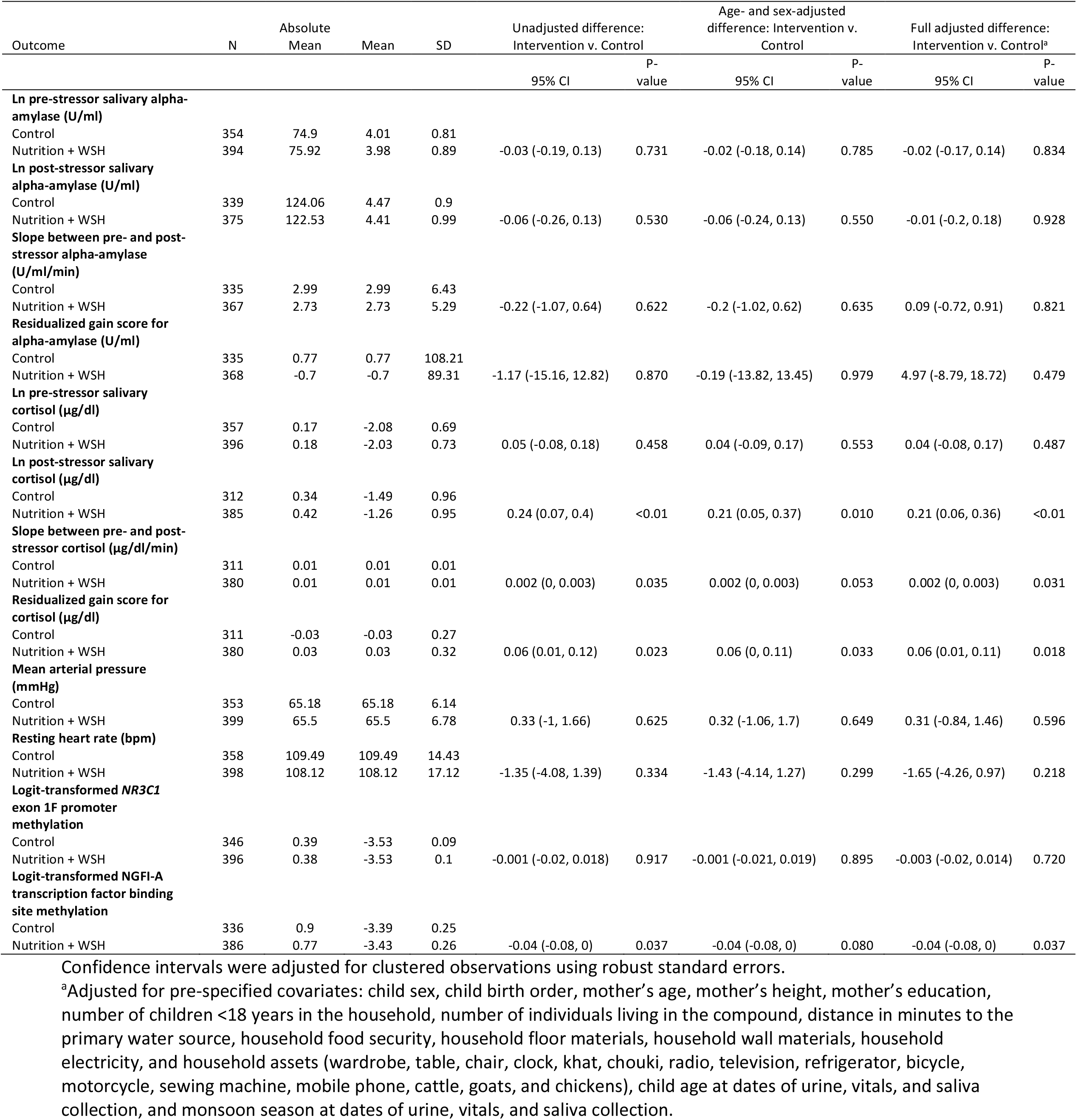
Effect of water, sanitation, handwashing, and nutrition intervention on stress response and DNA methylation measurements on Bangladeshi children at age 28 months.

A pre-specified subgroup analysis revealed that sex was not an effect modifier for F2-isoprostanes at year one (eTable 4 in Supplement). After two years, there was some evidence of effect measure modification with child sex: among males, the intervention group had a lower resting heart rate compared to the control group (−3.53 bpm, CI -6.62 to -0.44; *P*=0.025), and there was no effect among females (sex by treatment interaction *P*=0.027; eTable 5 in Supplement). Among females, the intervention group had higher pre-stressor cortisol levels compared to the control group (0.16 log μg/dl, CI 0.01 to 0.32; *P*=0.038), and there was no intervention effect on pre-stressor cortisol levels among males (sex by treatment interaction *P*=0.035).

## Discussion

In a setting with high levels of environmental contamination and food insecurity, the trial found that a combined drinking water, sanitation, handwashing, and nutritional intervention reduced oxidative stress, enhanced HPA axis functioning, and reduced methylation levels of the NGFI-A binding site in the *NR3C1* exon 1F promoter in young children. The magnitude of the effects of this environmental and nutritional intervention on cortisol production is within the range of intervention effects of psychosocial interventions reported in early childhood.^35^ The N+WSH intervention effects on F2-isoprostanes are also comparable to the effects of dietary interventions in adult populations.^36^

Oxidative stress, the accumulation of unstable free radicals that damage DNA and cellular structures, has been implicated in the pathophysiology of several pediatric disorders including asthma, protein-energy malnutrition, and diarrheal diseases.^37^ Augmented oxidative stress in children with severe forms of malnutrition, including kwashiorkor and marasmus, may be the consequence of increased production of reactive oxygen species or impaired antioxidant defenses.^38,39^ Micronutrients serve key roles in the body’s antioxidant defense system, either directly as antioxidants (e.g., vitamins C, A, and E) or indirectly as co-factors of antioxidant enzymes (e.g., manganese, copper, or zinc).^40^ The intervention reduced diarrhea, anemia, iron deficiency, and ultimately improved child growth.^28,41^ The lipid nutrient supplement containing ≥100% of the recommended daily allowance for 12 vitamins, including antioxidants such as vitamins C, A, and E and 9 minerals including manganese, copper, zinc, and selenium (full list available in Stewart et al.^41^) may have strengthened the antioxidant defenses of children in the intervention group compared to children in the control group. With enhanced antioxidant defenses, the bodies of children who received the intervention may have more efficiently scavenged free radicals, prevented their formation, and disrupted free-radical reactions.

As part of the innate immune response against pathogen invasion, phagocytic cells release reactive oxygen and nitrogen species that target proteins, DNA, and lipids.^42^ Innate immunity is tightly regulated because reactive oxygen and nitrogen species target pathogens and host cells alike. After one year, children in the intervention arm experienced less diarrhea and less enteric viral infections.^28,43^ It is plausible that the water, sanitation, and handwashing interventions interrupted pathogen transmission, leading to lower non-specific innate immune activity, less generation of reactive oxygen species, and reductions in downstream lipid peroxidation as measured by F2-isoprostanes. Together, less immune-activated production of reactive oxygen species and reinforced antioxidant defenses may have contributed to the lower levels of oxidative stress observed in children receiving the N+WSH interventions compared to children in the control group. These biological mechanisms could underpin the subsequent improvements in child development that we observed at age two years in the intervention group.^29^

The effects of the intervention were remarkably consistent across upstream and downstream levels of the HPA axis. Compared to the control group, the intervention group had hypomethylated NGFI-A transcription factor binding site, which leads to elevated glucocorticoid receptor gene expression.^44^ The higher post-stressor cortisol levels and cortisol reactivity exhibited in the intervention group are also indicative of increased activation of the glucocorticoid receptor. The mechanisms by which nutrition modulates the HPA axis have not been fully elucidated but could be mediated through the immune triad^15^ or epigenetic programming.^45^

In settings with inadequate water, sanitation, and hygiene infrastructure, infections are acquired early and frequently in childhood. Infections affect the transcriptional activation of the glucocorticoid receptor.^46^ In the WASH Benefits trial, children in arms receiving the WSH intervention had reduced *Giardia* and hookworm infections and lower acute respiratory illness at age two years compared to children in the control group.^47-49^ The hypomethylation of the NGFI-A transcription factor binding site and the robust cortisol response observed in the intervention group may have strengthened resistance to infections, as glucocorticoids are immunomodulatory hormones.^50^ During early childhood, a period of increased physiological plasticity and epigenetic programming, the combined water, sanitation, handwashing, and nutritional intervention enhanced children’s HPA axis regulatory capabilities, which in turn may have cascading effects on growth and development.

This study has limitations. One limitation of the study is that we only analyzed cortisol and sAA reactivity, which prevented us from characterizing the full cortisol and sAA awakening response, a common measure of HPA axis and autonomic nervous system functioning. Here, we report the effects of the combined intervention on stress response, because we did not also analyze samples from children who received only the nutrition and only the WSH intervention; thus, we cannot determine whether the effects were primarily driven by the nutrition intervention, the WSH intervention, or the combination of both. While selection bias is possible due to differential loss to follow-up, the IPCW results and the balanced household characteristics across both arms between children with outcome data and those lost to follow-up suggest that this type of bias was unlikely.

## Conclusions

In a low-resource setting, we found that an intensive combined drinking water, sanitation, handwashing, and nutritional intervention in early childhood reduced oxidative stress, enhanced the cortisol response, and reduced methylation levels of the glucocorticoid receptor gene. These findings support the future design and optimization of targeted nutritional and environmental therapeutic approaches that leverage physiologic plasticity to improve children’s health outcomes through the life course.

## Data Availability

Deidentified individual participant data collected for the study and a data dictionary defining each field in the set, will be made publicly available to others upon publication. The pre-specified, registered statistical analysis plan and replication files for the study will be available on Open Science Framework with publication (https://osf.io/9573v/).

## Acknowledgements

We greatly appreciate the families who participated in the study and the dedication of the icddr,b staff who delivered the interventions and collected the data and specimens.

## Funding/Support

This study was funded by Global Development grant OPPGD759 from the Bill & Melinda Gates Foundation to the University of California, Berkeley and by the National Institute of Allergy and Infectious Diseases of the National Institutes of Health [grant number K01AI136885 to A.L.]. icddr,b is grateful to the Governments of Bangladesh, Canada, Sweden, and the United Kingdom for providing core/unrestricted support. National Institute of Health (NIH) / National Cancer Institute (NCI) Comprehensive Cancer Center Grant [grant number P30-CA014236-47 to I.S.] provided support for the Duke Cancer Institute PK/PD Core Laboratory.

## Role of the Funder/Sponsor

The funders approved the study design, but were not involved in data collection, analysis, interpretation, or any decisions related to publication. The corresponding author had full access to all study data and final responsibility for the decision to submit for publication.

## Disclaimer

The content is solely the responsibility of the authors and does not necessarily represent the official views of the National Institutes of Health. In the interest of full disclosure, Douglas Granger is the founder and chief scientific and strategy advisor at Salimetrics LLC and SalivaBio LLC and these relationships are managed by the policies of the committees on conflict of interest at the Johns Hopkins University School of Medicine and the University of California at Irvine. Liying Yan is the president of EpigenDx, Inc. Ann Meyer is the Associate Director of Operations at EpigenDx, Inc.

## Data availability

Deidentified individual participant data collected for the study and a data dictionary defining each field in the set, will be made available to others. The pre-specified, registered statistical analysis plan and replication files for the study will be available on Open Science Framework with publication (https://osf.io/9573v/). The consort checklist for the study is included in the Supplement.

## Notes

### Clinical Trial

NCT01590095

### Author Declarations

Human subjects committees at the International Centre for Diarrhoeal Disease Research, Bangladesh, the University of California, Berkeley, and Stanford University gave ethical approval for this work.

