## Supplement for "Effects of drinking water, sanitation, handwashing and nutritional interventions on stress physiology, oxidative stress, and epigenetic programming in young children living in rural Bangladesh: A randomized clinical trial"

**Table of Contents**

**CONSORT Checklist**	2

**eMethods. Additional description**

Sample size calculations	4

Oxidative stress measurements	4

DNA extraction from saliva	4

Bisulfite treatment of gDNA and methylation analysis	4

References	5

**eFigures**

eFigure 1. Adjusted mean differences between the Control arm and the Nutrition + WSH arm for all

stress outcomes	6

eFigure 2. Unadjusted means and 95% confidence intervals by the Control arm and Nutrition + WSH

arms for all stress outcomes	7

**eTables**

eTable 1. Enrollment characteristics by intervention group within the WASH Benefits main trial

study population, within the stress study population enrolled in Year 1, and

within the stress study population lost to follow-up at Year 2	8

eTable 2. Effect of nutrition, water, sanitation, and handwashing intervention on oxidative stress

measurements on Bangladeshi children at age 14 months	9

eTable 3. Effect of nutrition, water, sanitation, and handwashing intervention on stress response

and DNA methylation measurements on Bangladeshi children at age 28 months	10

eTable 4. Effect modification with child sex on oxidative stress at age 14 months	11

eTable 5. Effect modification with child sex on stress response and DNA methylation

at age 28 months	12

**CONSORT Checklist**

| **Item** | **Description** | **Reported in Section** |
| --- | --- | --- |
| **Title and Abstract** | | |
| 1a | Identification as a randomized trial in the title; Identification as a cluster randomized trial in the title | Title |
| 1b | Structured summary of trial design, methods, results, and conclusions | Abstract |
| **Introduction** | | |
| Background and Objectives | | |
| 2a | Scientific background and explanation of rationale; Rationale for using a cluster design | Introduction |
| 2b | Specific objectives or hypotheses; Whether objectives pertain to the cluster level, the individual participant level, or both | Abstract; Introduction |
| **Methods** | | |
| Trial Design | | |
| 3a | Description of trial design (such as parallel, factorial) including allocation ratio; Definition of cluster and description of how the design features apply to the clusters | Methods |
| 3b | Important changes to methods after trial commencement (such as eligibility criteria), with reasons | N/A |
| Participants | | |
| 4a | Eligibility criteria for participants; Eligibility criteria for clusters | Methods (Participants) |
| 4b | Settings and locations where the data were collected | Methods |
| Interventions | | |
| 5 | The interventions for each group with sufficient details to allow replication, including how and when they were actually administered; Whether interventions pertain to the cluster level, the individual participant level, or both | Methods (Procedures) |
| Outcomes | | |
| 6a | Completely defined pre-specified primary and secondary outcome measures, including how and when they were assessed; Whether outcome measures pertain to the cluster level, the individual participant level, or both | Methods (Outcomes) |
| 6b | Any changes to trial outcomes after the trial commenced, with reasons | N/A |
| Sample Size | | |
| 7a | How sample size was determined; Method of calculation, number of cluster(s) (and whether equal or unequal cluster sizes are assumed), cluster size, a coefficient of intracluster correlation (ICC or k), and an indication of its uncertainty | Methods |
| 7b | When applicable, explanation of any interim analyses and stopping guidelines | N/A |
| **Randomization** | | |
| Sequence Generation | | |
| 8a | Method used to generate the random allocation sequence | Methods (Study design and randomization) |
| 8b | Type of randomization; details of any restriction (such as blocking and block size); Details of stratification or matching if used | Methods (Study design and randomization) |
| Allocation Concealment Mechanism | | |
| 9 | Mechanism used to implement the random allocation sequence (such as sequentially numbered containers), describing any steps taken to conceal the sequence until interventions were assigned; Specification that allocation was based on clusters rather than individuals and whether allocation concealment (if any) was at the cluster level, the individual participant level, or both | Methods (Study design and randomization) |
| Implementation | | |
| 10a | Who generated the random allocation sequence, who enrolled clusters, and who assigned clusters to interventions | Methods (Study design and randomization) |
| 10b | Mechanism by which individual participants were included in clusters for the purposes of the trial (such as complete enumeration, random sampling) | Methods (Study design and randomization) |
| 10c | From whom consent was sought (representatives of the cluster, or individual cluster members, or both) and whether consent was sought before or after randomization | Methods (Ethics) |
| Blinding | | |
| 11a | If done, who was blinded after assignment to interventions (for example, participants, care providers, those assessing outcomes)  and how | Methods (Study design and randomization) |
| 11b | If relevant, description of the similarity of interventions | N/A |
| Statistical Methods | | |
| 12a | Statistical methods used to compare groups for primary and secondary outcomes; How clustering was taken into account | Methods (Statistical analysis) |
| 12b | Methods for additional analyses, such as subgroup analyses and adjusted analyses | Methods (Statistical analysis) |
| **Results** | | |
| Participant Flow | | |
| 13a | For each group, the numbers of participants/clusters who were randomly assigned, received intended treatment, and were analyzed for the primary outcome | Results |
| 13b | For each group, losses and exclusions after randomization, together with reasons, for both clusters and individual cluster members | Methods (Participants) |
| Recruitment | | |
| 14a | Dates defining the periods of recruitment and follow-up | N/A |
| 14b | Why the trial ended or was stopped | N/A |
| Baseline Data | | |
| 15 | A table showing baseline demographic and clinical characteristics for each group; Baseline characteristics for the individual and cluster levels as applicable for each group | Table 1; eTable 1 in Supplement |
| Numbers Analyzed | | |
| 16 | For each group, number of participants/clusters (denominator) included in each analysis and whether the analysis was by the original assigned groups | Results; Table 1 |
| Outcomes and Estimation | | |
| 17a | For each primary and secondary outcome, results for each group, and the estimated effect size and its precision (such as 95% confidence interval); Results at the individual and cluster levels as applicable and a coefficient of intracluster correlation (ICC or k) for each primary outcome | Results; Tables 2 and 3; eTables 2-3 in Supplement |
| 17b | For binary outcome, presentation of both absolute and relative effect sizes is recommended | N/A |
| Ancillary Analyses | | |
| 18 | Results of any other analyses performed, including subgroup analyses and adjusted analyses, distinguishing pre-specified from exploratory | eTables 4-5 in Supplement |
| Harms | | |
| 19 | All important harms or unintended effects in each group | N/A |
| **Discussion** | | |
| Limitations | | |
| 20 | Trial limitations, addressing sources of potential bias, imprecision and, if relevant, multiplicity of analyses | Discussion |
| Generalizability | | |
| 21 | Generalizability (external validity, applicability) of the trial findings; Generalizability to clusters and/or individual participants (as relevant) | Discussion |
| Interpretation | | |
| 22 | Interpretation consistent with results, balancing benefits and harms, and considering other relevant evidence | Abstract; Discussion |
| **Other Information** | | |
| Registration | | |
| 23 | Registration number and name of trial registry | Methods (Statistical analysis) |
| Protocol | | |
| 24 | Where the full trial protocol can be accessed, if available | Methods (Statistical analysis) |
| Funding | | |
| 25 | Sources of funding and other support (such as supply of drugs), role of funders | Funding |

**Supplementary Methods**

*Sample size calculations*

Because the sample size calculations were based on the original environmental enteric dysfunction study,^1^ we assumed that this sample size would be sufficient to assess the stress response and DNA methylation outcomes of this substudy. The estimates for minimum detectable effects of nutrition and WSH interventions on these outcomes are outlined in the pre-registered analysis protocol.

*Oxidative stress measurements*

The F2-isoprostane isomers, iPF(2α)‐III, 2,3‐dinor‐iPF(2α)‐III, iPF(2α)‐VI, and 8,12‐iso‐iPF(2α)‐VI, were measured using liquid chromatography-tandem mass spectroscopy (LC-MS/MS) at Duke University as previously described and optimized for the present study.^2,3^ Urine creatinine (CR) concentration was measured to determine sample volume used for F2-isoprostane analysis. A larger urine volume (300 μL) was used in case of low CR (CR<0.6 mg/mL; highly diluted urine) to ensure assay sensitivity, a medium volume of urine (200 μL) was used when 0.6 mg/mL <CR<1 mg/mL, whereas a lower volume (100 μL) was used when CR was high (CR>1 mg/mL) to decrease matrix suppression effect on F2-isoprostane signals. To the appropriate volume of urine sample, 20 μL of 1M HCl, 20 μL of 100 ng/mL internal standard mix [iPF(2α)‐III‐d4, 8,12‐iso‐iPF(2α)‐VI‐d11, iPF(2α)‐VI‐d4], and 1 mL of methyltert-butylether (MTBE) was added and vigorously mixed in FastPrep (Thermo) for 3 x 45 sec at speed 4. After centrifugation, 800 μL of ether layer was evaporated (nitrogen stream), reconstituted in 50 μL methanol and 70 μL mobile phase A (see below) and 50 μL injected into Shimadzu 20A series / Applied Biosystems API 4000 QTrap LC/MS/MS instrument. Two C18 columns (Agilent Eclipse Plus, 150 × 4.6 mm and 50 x 4.6 mm, 1.8 µm) in series were used with 0.1% acetic acid as mobile phase A and methanol as mobile phase B delivered as 40-75% B gradient elution over 26 min. The mass spectrometer was operated in negative mode with the following MS/MS transitions (m/z): 353/193 [iPF(2α)‐III], 357/197 [iPF(2α)‐III‐d4], 325/237 [2,3‐dinor‐iPF(2α)‐III], 353/115 [iPF(2α)‐VI and 8,12‐iso‐iPF(2α)‐VI], 364/115 [iPF(2α)‐VI‐d11], and 357/115 [8,12‐iso‐iPF(2α)‐VI‐d4]. Lower limits of quantification (LLOQ >80% accuracy) were 0.063, 0.31, 0.63, and 0.63 mg/mL for iPF(2α)‐III, 2,3‐dinor‐iPF(2α)‐III, iPF(2α)‐VI, and 8,12‐iso‐iPF(2α)‐VI, respectively. The concentration of F2‐isoprostanes was adjusted for urinary creatinine (CR) to account for urine diluteness. Creatinine (CR) was measured after 1/1000 dilution of urine by deionized water, centrifugation, and direct injection into LC/MS/MS system. Agilent Eclipse Plus 50 x 4.6 mm, 1.8 μm column was used for separation. CR and CR-d3 (internal standard) were measured at m/z=114/44 and m/z=117/47, respectively.

*DNA extraction from saliva*

Saliva samples were collected using the Oragene kit (OGR-575). DNA was extracted from 200 µL saliva using DNAdvance (Beckmen Coulter) with the Biomek FXP liquid handler (Beckman Coulter) at EpigenDx (Hopkinton, MA). The NanoDrop 2000 (ThermoFisher Scientific) was used to quantify the extracted DNA by OD260/280.

*Bisulfite treatment of gDNA and methylation analysis*

EpigenDx carried out pyrosequencing of bisulfite-treated DNA. Briefly, 500 ng of extracted genomic DNA was bisulfite treated using the EZ DNA Methylation kit (Zymo Research, Inc., CA). The kit protocol was followed for purification and elution of the bisulfite treated DNA (final elution volume of 46 µL). PCR amplification was achieved using 1 µL of bisulfite treated DNA and 0.2 µM of each primer. To purify the final PCR product using sepharose beads, one primer was biotin-labeled and purified by high performance liquid chromatography.

After being bound to Streptavidin Sepharose HP (GE Healthcare Life Sciences), the immobilized PCR products were purified, washed, denatured with a 0.2 µM NaOH solution, and rewashed using the Pyrosequencing Vacuum Prep Tool (Pyrosequencing, Qiagen), according to the manufacturer’s instructions. The *NR3C1* pyrosequencing methylation assay target region is listed below:

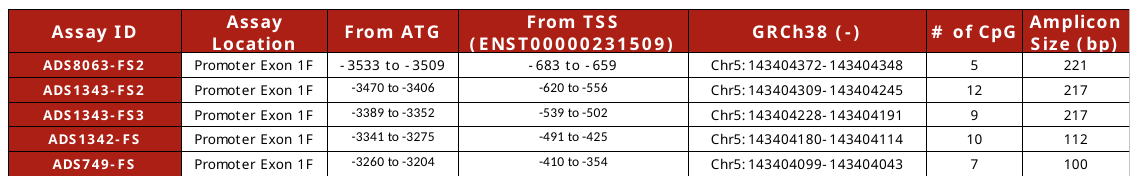

Purified single stranded PCR products were annealed to 0.5 µM of sequencing primer. Following the manufacturer’s protocol, 10 µL of the PCR products were pyrosequenced on the PSQ96 HS System (Pyrosequencing, Qiagen). QCpG software (Pyrosequencing, Qiagen) was used to analyze the methylation status of each locus (CpG site) individually as an artificial C/T SNP. To calculate the methylation level at each CpG site, the following formula was used: the percentage of methylated alleles divided by the sum of all methylated and unmethylated alleles. To obtain the mean methylation level, the methylation levels of all measured CpG sites within the targeted region of the gene were used. To ensure detection of incomplete bisulfite conversion of the DNA, each experiment used non-CpG cytosines as internal controls. Other controls in each PCR included unmethylated and methylated DNA. To test for bias, unmethylated control DNA was combined with *in vitro* methylated DNA at several ratios (0%, 5%, 10%, 25%, 50%, 75%, and 100%), the mixed products were bisulfite-modified and underwent PCR, followed by pyrosequencing analysis.

*References*

1.	Lin A, Ali S, Arnold BF, et al. Effects of water, sanitation, handwashing, and nutritional interventions on environmental enteric dysfunction in young children: a cluster-randomized controlled trial in rural Bangladesh. *Clin Infect Dis.* 2019.

2.	Il'yasova D, Spasojevic I, Wang F, et al. Urinary biomarkers of oxidative status in a clinical model of oxidative assault. *Cancer Epidemiol Biomarkers Prev.* 2010;19(6):1506-1510.

3.	Zhang H, Il'yasova D, Sztaray J, Young SP, Wang F, Millington DS. Quantification of the oxidative damage biomarker 2,3-dinor-8-isoprostaglandin-F(2alpha) in human urine using liquid chromatography-tandem mass spectrometry. *Anal Biochem.* 2010;399(2):302-304.

**eFigure 1. Adjusted mean differences between the Control arm and the Nutrition + WSH arm for all stress outcomes**

**eFigure 2. Unadjusted means and 95% confidence intervals by the Control arm and Nutrition + WSH arms for all stress outcomes**

**eTable 1. Enrollment characteristics by intervention group within the WASH Benefits main trial study population, within the stress study population enrolled in Year 1, and within the stress study population lost to follow-up at Year 2**

|  | WASH Benefits Main Trial | | Stress Status Study: Had outcomes at Year 1 | | Stress Status Study: Lost to follow-up at Year 2 | |
| --- | --- | --- | --- | --- | --- | --- |
| No. of compounds: | Control (N=1382) | N + WSH Intervention (N=686) | Control (N=332) | N+WSH Intervention (N=356) | Control (N=45) | N + WSH Intervention (N=38) |
| **Maternal** |  |  |  |  |  |  |
| Age(years) | 24 (5) | 24 (6) | 23 (5) | 24 (6) | 23 (5) | 24 (6) |
| Years of education | 6 (3) | 6 (3) | 7 (3) | 6 (3) | 7 (3) | 5 (4) |
| **Paternal** |  |  |  |  |  |  |
| Years of education | 5 (4) | 5 (4) | 6 (4) | 5 (4) | 6 (3) | 5 (4) |
| Works in agriculture | 414 (30%) | 207 (30%) | 80 (24%) | 101 (28%) | 10 (22%) | 9 (24%) |
| **Household** |  |  |  |  |  |  |
| Number of people | 5 (2) | 5 (2) | 5 (3) | 5 (2) | 5 (3) | 5 (2) |
| Has electricity | 784 (57%) | 412 (60%) | 200 (60%) | 214 (60%) | 27 (60%) | 20 (53%) |
| Has a cement floor | 145 (10%) | 72 (10%) | 59 (18%) | 43 (12%) | 8 (18%) | 2 (5%) |
| Acres of agricultural land owned | 0.15 (0.21) | 0.14 (0.38) | 0.18 (0.25) | 0.13 (0.13) | 0.18 (0.25) | 0.09 (0.09) |
| **Drinking Water** |  |  |  |  |  |  |
| Shallow tubewell primary water source | 1038 (75%) | 504 (73%) | 236 (71%) | 249 (70%) | 31 (69%) | 24 (63%) |
| Stored water observed at home | 666 (48%) | 331 (48%) | 168 (51%) | 185 (52%) | 27 (60%) | 18 (47%) |
| Reported treating water yesterday | 4 (0%) | 2 (0%) | 1 (0%) | 1 (0%) | 0 (0%) | 0 (0%) |
| Distance (mins) to primary water source | 1 (3) | 1 (2) | 1 (2) | 1 (2) | 1 (1) | 1 (1) |
| **Sanitation** |  |  |  |  |  |  |
| Reported daily open defecation |  |  |  |  |  |  |
| Adult men | 97 (7%) | 50 (7%) | 12 (4%) | 31 (9%) | 0 (0%) | 3 (8%) |
| Adult women | 62 (4%) | 24 (4%) | 8 (2%) | 16 (5%) | 0 (0%) | 1 (3%) |
| Children: 8 to <15 years | 53 (10%) | 28 (10%) | 6 (4%) | 18 (11%) | 2 (10%) | 3 (20%) |
| Children: 3 to <8 years | 267 (38%) | 134 (37%) | 47 (29%) | 68 (33%) | 8 (42%) | 7 (28%) |
| Children: 0 to <3 years^a^ | 245 (82%) | 123 (88%) | 55 (71%) | 69 (88%) | 6 (67%) | 6 (86%) |
| Latrine |  |  |  |  |  |  |
| Owned^b^ | 750 (54%) | 367 (53%) | 208 (63%) | 191 (54%) | 33 (73%) | 18 (47%) |
| Concrete Slab | 1251 (95%) | 621 (94%) | 315 (97%) | 317 (93%) | 45 (100%) | 37 (100%) |
| Functional water seal | 358 (31%) | 155 (27%) | 118 (39%) | 89 (32%) | 21 (48%) | 11 (35%) |
| Visible stool on slab or floor | 625 (48%) | 298 (46%) | 149 (46%) | 177 (53%) | 17 (38%) | 20 (54%) |
| Owned a child potty | 61 (4%) | 30 (4%) | 27 (8%) | 16 (4%) | 9 (20%) | 0 (0%) |
| Human feces observed in the |  |  |  |  |  |  |
| House | 114 (8%) | 49 (8%) | 23 (7%) | 30 (8%) | 5 (11%) | 2 (5%) |
| Child's play area | 21 (2%) | 7 (1%) | 4 (1%) | 6 (2%) | 1 (2%) | 0 (0%) |
| **Handwashing location** |  |  |  |  |  |  |
| Within six steps of latrine |  |  |  |  |  |  |
| Has water | 178 (14%) | 72 (11%) | 57 (19%) | 45 (14%) | 5 (12%) | 2 (6%) |
| Has soap | 88 (7%) | 36 (6%) | 30 (10%) | 20 (6%) | 4 (10%) | 2 (6%) |
| Within six steps of kitchen |  |  |  |  |  |  |
| Has water | 118 (9%) | 60 (9%) | 32 (10%) | 33 (10%) | 2 (5%) | 2 (6%) |
| Has soap | 33 (3%) | 18 (3%) | 12 (4%) | 9 (3%) | 0 (0%) | 0 (0%) |
| **Nutrition** |  |  |  |  |  |  |
| Household is food secure^c^ | 932 (67%) | 485 (71%) | 246 (74%) | 251 (71%) | 34 (76%) | 25 (66%) |

Data are n (%) or mean (SD). Percentages were estimated from slightly smaller denominators than those shown at the top of the table for the following variables due to missing values: mother’s age, father’s education, father works in agriculture, acres of land owned, open defecation, latrine has a concrete slab, latrine has a functional water seal, visible stool on latrine slab or floor, ownership of child potty, observed feces in the house or child’s play area, handwashing variables.

^a^Open defecation does not include diaper disposal of feces.

^b^Households who do not own a latrine typically share a latrine with extended family members who live in the same compound.

^c^Assessed by the Household Food Insecurity Access Scale.

**eTable 2. Effect of nutrition, water, sanitation, and handwashing intervention on oxidative stress measurements on Bangladeshi children at age 14 months**

| Urinary  F2-isoprostanes | N | Absolute Mean | Mean | SD | Unadjusted difference: Intervention v. Control | | Age- and sex-adjusted difference: Intervention v. Control | | Fully adjusted difference: Intervention v. Control^a^ | | IPCW adjusted difference: Intervention v. Control^b^ | |
| --- | --- | --- | --- | --- | --- | --- | --- | --- | --- | --- | --- | --- |
|  |  |  |  |  | 95% CI | P-value | 95% CI | P-value | 95% CI | P-value | 95% CI | P-value |
| **Ln iPF(2α)-III (ng/mg creatinine)** |  |  |  |  |  |  |  |  |  |  |  |  |
| Control | 332 | 0.83 | -0.33 | 0.54 |  |  |  |  |  |  |  |  |
| Nutrition + WSH | 356 | 0.7 | -0.5 | 0.56 | -0.16 (-0.27, -0.06) | <0.01 | -0.17 (-0.28, -0.06) | <0.01 | -0.14 (-0.22, -0.05) | <0.01 | -0.14 (-0.17, -0.11) | <0.001 |
| **Ln 2,3-dinor-iPF(2α)-III (ng/mg creatinine)** |  |  |  |  |  |  |  |  |  |  |  |  |
| Control | 332 | 6.66 | 1.84 | 0.33 |  |  |  |  |  |  |  |  |
| Nutrition + WSH | 356 | 5.64 | 1.68 | 0.32 | -0.16 (-0.23, -0.09) | <0.001 | -0.19 (-0.26, -0.13) | <0.001 | -0.18 (-0.24, -0.12) | <0.001 | -0.22 (-0.24, -0.2) | <0.001 |
| **Ln iPF(2α)-VI (ng/mg creatinine)** |  |  |  |  |  |  |  |  |  |  |  |  |
| Control | 332 | 16.36 | 2.68 | 0.46 |  |  |  |  |  |  |  |  |
| Nutrition + WSH | 356 | 13.5 | 2.5 | 0.44 | -0.17 (-0.25, -0.1) | <0.001 | -0.2 (-0.27, -0.13) | <0.001 | -0.19 (-0.25, -0.12) | <0.001 | -0.11 (-0.14, -0.08) | <0.001 |
| **Ln 8,12-iso-iPF(2α)-VI (ng/mg creatinine)** |  |  |  |  |  |  |  |  |  |  |  |  |
| Control | 332 | 16.05 | 2.63 | 0.55 |  |  |  |  |  |  |  |  |
| Nutrition + WSH | 356 | 13.49 | 2.44 | 0.57 | -0.19 (-0.29, -0.1) | <0.001 | -0.21 (-0.31, -0.12) | <0.001 | -0.24 (-0.32, -0.16) | <0.001 | -0.18 (-0.22, -0.15) | <0.001 |

Confidence intervals were adjusted for clustered observations using robust standard errors.

^a^Adjusted for pre-specified covariates: child sex, child birth order, mother’s age, mother’s height, mother’s education, number of children <18 years in the household, number of individuals living in the compound, distance in minutes to the primary water source, household food security, household floor materials, household wall materials, household electricity, and household assets (wardrobe, table, chair, clock, khat, chouki, radio, television, refrigerator, bicycle, motorcycle, sewing machine, mobile phone, cattle, goats, and chickens), child age at dates of urine, vitals, and saliva collection, and monsoon season at dates of urine, vitals, and saliva collection.

^b^Inverse probability of censoring weighting.

**eTable 3. Effect of nutrition, water, sanitation, and handwashing intervention on stress response and DNA methylation measurements on Bangladeshi children at age 28 months**

| Outcome | N | Absolute Mean | Mean | SD | Unadjusted difference: Intervention v. Control | | Age- and sex-adjusted difference: Intervention v. Control | | Fully adjusted difference: Intervention v. Control^a^ | | IPCW adjusted difference: Intervention v. Control^b^ | |
| --- | --- | --- | --- | --- | --- | --- | --- | --- | --- | --- | --- | --- |
|  |  |  |  |  | 95% CI | P-value | 95% CI | P-value | 95% CI | P-value | 95% CI | P-value |
| **Ln pre-stressor Salivary alpha-amylase (U/ml)** |  |  |  |  |  |  |  |  |  |  |  |  |
| Control | 354 | 74.9 | 4.01 | 0.81 |  |  |  |  |  |  |  |  |
| Nutrition + WSH | 394 | 75.92 | 3.98 | 0.89 | -0.03 (-0.19, 0.13) | 0.731 | -0.02 (-0.18, 0.14) | 0.785 | -0.02 (-0.17, 0.14) | 0.834 | -0.01 (-0.15, 0.13) | 0.914 |
| **Ln post-stressor Salivary alpha-amylase (U/ml)** |  |  |  |  |  |  |  |  |  |  |  |  |
| Control | 339 | 124.06 | 4.47 | 0.9 |  |  |  |  |  |  |  |  |
| Nutrition + WSH | 375 | 122.53 | 4.41 | 0.99 | -0.06 (-0.26, 0.13) | 0.530 | -0.06 (-0.24, 0.13) | 0.550 | -0.01 (-0.2, 0.18) | 0.928 | -0.02 (-0.08, 0.04) | 0.470 |
| **Slope between pre- and post-stressor alpha-amylase (U/ml/min)** |  |  |  |  |  |  |  |  |  |  |  |  |
| Control | 335 | 2.99 | 2.99 | 6.43 |  |  |  |  |  |  |  |  |
| Nutrition + WSH | 367 | 2.73 | 2.73 | 5.29 | -0.22 (-1.07, 0.64) | 0.622 | -0.2 (-1.02, 0.62) | 0.635 | 0.09 (-0.72, 0.91) | 0.821 | 0.3 (-0.66, 1.26) | 0.542 |
| **Residualized gain score for alpha-amylase (U/ml)** |  |  |  |  |  |  |  |  |  |  |  |  |
| Control | 335 | 0.77 | 0.77 | 108.21 |  |  |  |  |  |  |  |  |
| Nutrition + WSH | 368 | -0.7 | -0.7 | 89.31 | -1.17 (-15.16, 12.82) | 0.870 | -0.19 (-13.82, 13.45) | 0.979 | 4.97 (-8.79, 18.72) | 0.479 | 17.77 (12.08, 23.47) | <0.001 |
| **Ln pre-stressor salivary cortisol (µg/dl)** |  |  |  |  |  |  |  |  |  |  |  |  |
| Control | 357 | 0.17 | -2.08 | 0.69 |  |  |  |  |  |  |  |  |
| Nutrition + WSH | 396 | 0.18 | -2.03 | 0.73 | 0.05 (-0.08, 0.18) | 0.458 | 0.04 (-0.09, 0.17) | 0.553 | 0.04 (-0.08, 0.17) | 0.487 | 0.05 (0.01, 0.09) | <0.01 |
| **Ln post-stressor salivary cortisol (µg/dl)** |  |  |  |  |  |  |  |  |  |  |  |  |
| Control | 312 | 0.34 | -1.49 | 0.96 |  |  |  |  |  |  |  |  |
| Nutrition + WSH | 385 | 0.42 | -1.26 | 0.95 | 0.24 (0.07, 0.4) | <0.01 | 0.21 (0.05, 0.37) | 0.010 | 0.21 (0.06, 0.36) | <0.01 | 0.2 (0.06, 0.34) | <0.01 |
| **Slope between pre- and post-stressor cortisol (µg/dl/min)** |  |  |  |  |  |  |  |  |  |  |  |  |
| Control | 311 | 0.01 | 0.01 | 0.01 |  |  |  |  |  |  |  |  |
| Nutrition + WSH | 380 | 0.01 | 0.01 | 0.01 | 0.002 (0, 0.003) | 0.035 | 0.002 (0, 0.003) | 0.053 | 0.002 (0, 0.003) | 0.031 | 0.003 (0.001, 0.004) | <0.001 |
| **Residualized gain score for cortisol (µg/dl)** |  |  |  |  |  |  |  |  |  |  |  |  |
| Control | 311 | -0.03 | -0.03 | 0.27 |  |  |  |  |  |  |  |  |
| Nutrition + WSH | 380 | 0.03 | 0.03 | 0.32 | 0.06 (0.01, 0.12) | 0.023 | 0.06 (0, 0.11) | 0.033 | 0.06 (0.01, 0.11) | 0.018 | 0.09 (0.04, 0.15) | <0.01 |
| **Mean arterial pressure (mmHg)** |  |  |  |  |  |  |  |  |  |  |  |  |
| Control | 353 | 65.18 | 65.18 | 6.14 |  |  |  |  |  |  |  |  |
| Nutrition + WSH | 399 | 65.5 | 65.5 | 6.78 | 0.33 (-1, 1.66) | 0.625 | 0.32 (-1.06, 1.7) | 0.649 | 0.31 (-0.84, 1.46) | 0.596 | 0.46 (0.06, 0.86) | 0.025 |
| **Resting heart rate (bpm)** |  |  |  |  |  |  |  |  |  |  |  |  |
| Control | 358 | 109.49 | 109.49 | 14.43 |  |  |  |  |  |  |  |  |
| Nutrition + WSH | 398 | 108.12 | 108.12 | 17.12 | -1.35 (-4.08, 1.39) | 0.334 | -1.43 (-4.14, 1.27) | 0.299 | -1.65 (-4.26, 0.97) | 0.218 | -1.36 (-2.29, -0.42) | <0.01 |
| **Logit-transformed *NR3C1* exon 1F promoter methylation** |  |  |  |  |  |  |  |  |  |  |  |  |
| Control | 346 | 0.39 | -3.53 | 0.09 |  |  |  |  |  |  |  |  |
| Nutrition + WSH | 396 | 0.38 | -3.53 | 0.1 | -0.001 (-0.02, 0.018) | 0.917 | -0.001 (-0.021, 0.019) | 0.895 | -0.003 (-0.02, 0.014) | 0.720 | -0.01 (-0.02, 0) | <0.01 |
| **Logit-transformed NGFI-A transcription factor binding site methylation** |  |  |  |  |  |  |  |  |  |  |  |  |
| Control | 336 | 0.9 | -3.39 | 0.25 |  |  |  |  |  |  |  |  |
| Nutrition + WSH | 386 | 0.77 | -3.43 | 0.26 | -0.04 (-0.08, 0) | 0.037 | -0.04 (-0.08, 0) | 0.080 | -0.04 (-0.08, 0) | 0.037 | -0.05 (-0.09, -0.01) | 0.017 |

Confidence intervals were adjusted for clustered observations using robust standard errors.

^a^Adjusted for pre-specified covariates: child sex, child birth order, mother’s age, mother’s height, mother’s education, number of children <18 years in the household, number of individuals living in the compound, distance in minutes to the primary water source, household food security, household floor materials, household wall materials, household electricity, and household assets (wardrobe, table, chair, clock, khat, chouki, radio, television, refrigerator, bicycle, motorcycle, sewing machine, mobile phone, cattle, goats, and chickens), child age at dates of urine, vitals, and saliva collection, and monsoon season at dates of urine, vitals, and saliva collection.

^b^Inverse probability of censoring weighting.

**eTable 4. Effect modification with sex at 14 months**

|  | Female | | | | | Male | | | | | Interaction P-value |
| --- | --- | --- | --- | --- | --- | --- | --- | --- | --- | --- | --- |
| Urinary F2-isoprostanes | N | Mean | SD | Unadjusted difference: Intervention vs. Control (95% CI) | P-value | N | Mean | SD | Unadjusted difference: Intervention vs. Control (95% CI) | P-value |  |
| Ln iPF(2α)-III (ng/mg creatinine) | 349 | 0.77 | 0.42 | -0.11 (-0.23, 0) | 0.053 | 339 | 0.75 | 0.56 | -0.22 (-0.35, -0.1) | <0.001 | 0.231 |
| Ln 2,3-dinor-iPF(2α)-III (ng/mg creatinine) | 349 | 6.33 | 2.37 | -0.21 (-0.29, -0.13) | <0.001 | 339 | 5.94 | 1.92 | -0.12 (-0.19, -0.05) | <0.01 | 0.051 |
| Ln iPF(2α)-VI (ng/mg creatinine) | 349 | 15.3 | 7.28 | -0.16 (-0.26, -0.06) | <0.01 | 339 | 14.44 | 11.73 | -0.19 (-0.29, -0.09) | <0.001 | 0.712 |
| Ln 8,12-iso-iPF(2α)-VI (ng/mg creatinine) | 349 | 15.66 | 9.65 | -0.22 (-0.34, -0.09) | <0.001 | 339 | 13.77 | 7.93 | -0.18 (-0.3, -0.06) | <0.01 | 0.628 |

**eTable 5. Effect modification with sex at 28 months**

|  | Female | | | | | Male | | | | | | Interaction P-value |
| --- | --- | --- | --- | --- | --- | --- | --- | --- | --- | --- | --- | --- |
| Outcome | N | Mean | SD | Unadjusted difference: Intervention vs. Control (95% CI) | P-value | N | Mean | SD | Unadjusted difference: Intervention vs. Control (95% CI) | P-value |  | |
| Mean arterial pressure (mmHg) | 393 | 65.26 | 6.45 | -0.01 (-1.43, 1.41) | 0.993 | 359 | 65.45 | 6.53 | 0.67 (-0.97, 2.3) | 0.427 | 0.496 | |
| Resting heart rate (bpm) | 393 | 110.15 | 16.24 | 0.57 (-2.3, 3.44) | 0.697 | 363 | 107.27 | 15.42 | -3.53 (-6.62, -0.44) | 0.025 | 0.027 | |
| Ln pre-stressor alpha-amylase (U/ml) | 388 | 70.87 | 56.22 | -0.03 (-0.2, 0.14) | 0.718 | 360 | 80.35 | 68.76 | -0.02 (-0.19, 0.15) | 0.804 | 0.920 | |
| Ln post-stressor alpha-amylase (U/ml) | 371 | 122.57 | 126.06 | -0.02 (-0.21, 0.17) | 0.873 | 343 | 124 | 97.8 | -0.11 (-0.33, 0.11) | 0.328 | 0.536 | |
| Ln pre-stressor cortisol (µg/dl) | 390 | 0.19 | 0.24 | 0.16 (0.01, 0.32) | 0.038 | 363 | 0.16 | 0.13 | -0.08 (-0.23, 0.08) | 0.330 | 0.035 | |
| Ln post-stressor cortisol (µg/dl) | 363 | 0.4 | 0.38 | 0.31 (0.14, 0.47) | <0.001 | 334 | 0.37 | 0.31 | 0.15 (-0.04, 0.34) | 0.128 | 0.188 | |
| Logit-transformed *NR3C1* exon 1F promoter methylation | 387 | 0.4 | 0.35 | 0 (-0.02, 0.03) | 0.708 | 355 | 0.36 | 0.25 | -0.01 (-0.03, 0.01) | 0.425 | 0.345 | |
| Logit-transformed NGFI-A transcription factor binding site methylation | 379 | 0.82 | 0.94 | -0.02 (-0.07, 0.03) | 0.414 | 343 | 0.85 | 0.96 | -0.07 (-0.12, -0.01) | 0.012 | 0.166 | |
| Slope between pre- and post-stressor alpha-amylase (U/ml/min) | 364 | 3.02 | 6.65 | 0.23 (-1.16, 1.63) | 0.741 | 338 | 2.67 | 4.87 | -0.81 (-2, 0.39) | 0.186 | 0.277 | |
| Slope between pre- and post-stressor cortisol (µg/dl/min) | 358 | 0.01 | 0.01 | 0 (0, 0) | 0.052 | 333 | 0.01 | 0.01 | 0 (0, 0) | 0.079 | 0.862 | |
| Residualized gain score for alpha-amylase (U/ml) | 364 | 3.15 | 113.43 | 5.12 (-18.6, 28.84) | 0.672 | 339 | -3.38 | 79.94 | -8.7 (-26.76, 9.35) | 0.345 | 0.380 | |
| Residualized gain score for cortisol (µg/dl) | 358 | 0 | 0.32 | 0.07 (0, 0.13) | 0.036 | 333 | 0 | 0.28 | 0.06 (0, 0.12) | 0.063 | 0.774 | |
